# Structural plasticity following deep brain stimulation of the internal capsule in treatment resistant depression

**DOI:** 10.1101/2024.12.12.24318914

**Authors:** Z. Zhong, I.O. Bergfeld, B.P. de Kwaasteniet, J. Luigjes, R.J.T. Mocking, J. van Laarhoven, P. Notten, G. Beute, P. van den Munckhof, P.R. Schuurman, D.A.J.P. Denys, G.A. van Wingen

## Abstract

**Objective:** Deep brain stimulation (DBS) targeting the ventral anterior limb of the internal capsule (vALIC) shows potential as treatment for treatment resistant depression (TRD). While DBS alters brain function, it is not yet known whether it also induces anatomical alterations. Here we investigated the long-term effects of vALIC DBS on brain structure using structural magnetic resonance imaging.

**Methods:** We included data from twelve patients with TRD before DBS surgery and after DBS parameter optimization, and from sixteen matched healthy controls at a comparable time interval to control for test-retest effects. To investigate the short-term effects of DBS deactivation after parameter optimization, thirteen patients were additionally scanned after double-blind periods of active and sham stimulation. Voxel-based morphometry analysis was used to identify volumetric differences.

**Result:** The group x time interaction showed significant changes in anterior thalamic gray matter volume, with a relative reduction in TRD patients compared to controls. Follow-up analysis suggested that this was related to larger thalamic volume at baseline in DBS non-responders. A direct comparison between responders and non-responders showed an additional interaction in the posterior thalamus and hippocampus, which was also related to larger volumes at baseline in non-responders. The comparison between active and sham stimulation during the crossover phase did not show significant differences.

**Conclusion:** These results show that long-term vALIC DBS is associated with a reduction in thalamic volume compared to healthy controls, suggesting that long-term DBS induces focal structural plasticity.

## Introduction

It is estimated that up to one third of patients with major depression disorder (MDD) do not sufficiently respond to conventional antidepressant treatments, including medication, psychotherapy and electroconvulsive therapy (ECT)([1). Deep brain stimulation (DBS) is an emerging treatment for patients with treatment-resistant depression (TRD), which involves electrical stimulation of brain regions such as the subcallosal cingulate gyrus, ventral striatum/ventral capsule, nucleus accumbens, inferior thalamic peduncle, medial forebrain bundle and ventral anterior limb of the internal capsule (vALIC)(2). Open-label studies frequently indicate a reduction in depression severity over extended periods of stimulation (3).

We found that DBS of the vALIC, which consists of white matter bundles that connect the prefrontal cortex with the thalamus and midbrain, improves depressive symptoms in TRD patients (4). Additionally, active DBS was significantly more effective than sham DBS (5). Similar effects have been reported when targeting the superolateral medial forebrain bundle (slMFB) (6). This is one of the white matter bundles that travels through the ALIC, and was tested as a new target due to its putative dysfunction in TRD and key function in the human reward system (7). The other main white matter bundle in the ALIC, the anterior thalamic radiation (ATR), may be more involved in negative emotions (8). Thus, although we know which white matter bundles are targeted, it still remains unclear how vALIC-DBS affects the brain regions they connect.

Previous neuroimaging studies have explored the possible mechanisms of different targets of DBS in TRD. Stimulation in the subcallosal cingulate cortex (SCC) or the ventral capsule/ventral striatum (VC/VS) has been shown to decrease activity in the SCC and the nucleus accumbens accumbens (NAc) (9-11), while another study found increased activity in these areas after DBS (12). Positive clinical response to SCC DBS is associated with decreased activity in the SCC, medial prefrontal cortex (mPFC), orbitofrontal cortex (OFC), and insula following chronic stimulation (9, 12). Stimulation of the vALI restores right amygdala responsivity (13). However, since this all concerns functional brain changes, it remains unknown whether DBS also leads to structural brain changes in TRD patients.

In this study, we used voxel-based morphometry (VBM) to analyze structural magnetic resonance imaging (sMRI) scans from TRD patients before vALIC DBS implantation and after DBS parameter optimization. Matched healthy controls were included and received sMRI at two comparable time points to control for naturalistic changes in brain structure. Besides the structural changes due to electrode implantation, we hypothesized that structural plasticity may primarily occur within the white matter target bundles (anterior thalamic radiation), its gray matter connections (thalamus and prefrontal cortex), and the brain regions that show the largest structural plasticity after electroconvulsive therapy (hippocampus and amygdala) (14). In addition, we assessed short-term effects of vALIC DBS deactivation after parameter optimization by analyzing the sMRI scans that were obtained during double-blind periods of active and sham stimulation.

## METHODS

### Patient selection

In our longitudinal study, we enrolled patients with TRD and healthy controls between March 2010 and August 2016. Patients underwent a randomized crossover phase, which was not applicable to the healthy controls. This imaging study was conducted as an adjunct to a previously published clinical trial investigating DBS for TRD (trial registration number: NL-OMON23670). Approval for the study was obtained from the medical ethics committees at both participating hospitals: the Academic Medical Center in Amsterdam [AMC] and St. Elisabeth Hospital in Tilburg [ETZ]. All participants provided written informed consent before participating in the study.

Upon enrollment, patients were required to meet the following criteria: aged between 18 and 65 years, diagnosed with MDD according to the DSM-IV criteria, with an illness duration exceeding 2 years, a 17-item Hamilton Depression Rating Scale (HAM-D-17) score of 18 or higher, and a Global Assessment of Function (GAF) score of 45 or lower. Additionally, patients had to demonstrate treatment resistance, defined as failure to respond to at least two different classes of second-generation antidepressants, one trial of a tricyclic antidepressant, one trial of a tricyclic antidepressant with lithium augmentation, one trial of a monoamine oxidase inhibitor, and at least six sessions of bilateral ECT. Patients who met these criteria and maintained stability with maintenance ECT but relapsed upon its discontinuation were also considered eligible. Furthermore, patients needed to possess the cognitive capacity (IQ > 80) to comprehend the procedure’s implications and make decisions independently. Exclusion criteria encompassed Parkinson’s disease, dementia, epilepsy, bipolar disorder, schizophrenia or a history of psychosis unrelated to MDD, antisocial personality disorder, current tic disorder, depression attributed to an organic cause, substance abuse within the past 6 months, unstable physical condition, pregnancy, or general contraindications for surgery.

Healthy controls were matched by age, sex, and education level. They and their first-degree relatives needed to have negative lifetime histories of psychiatric illness.

### DBS Treatment

Bilateral implantation of four-contact leads was performed, with connections made to a neurostimulator (lead: 3389; stimulator: Activa PC, Medtronic, Minneapolis, MN, USA). The electrodes were positioned such that the most ventral contact targeted the core of the NAc. The three more dorsal contacts were placed within the vALIC. Following a recovery period of three weeks, optimization of the DBS settings commenced. Optimization concluded once a stable response was achieved for a minimum of four weeks or after a maximum duration of 52 weeks. Further details regarding the surgical procedure and DBS treatment can be found in previous publications (5).

### Study Design

MRI scans were conducted three weeks prior to DBS surgery (baseline) and after the optimization of DBS parameters (follow-up). Efforts were made to maintain stability in concurrent medication during the optimization phase, though psychiatrists retained the discretion to make adjustments based on clinical necessity (for a comprehensive overview of psychotropic medications administered over time, refer to Supplementary Table 1). Healthy controls underwent MRI scans at two distinct time points (baseline and after a 5-month follow-up). Subsequent to the follow-up assessments, participants in the TRD group proceeded to the double-blind randomized crossover phase. This phase comprised two blocks lasting one to six weeks each, during which DBS stimulation was either activated (active) or deactivated (sham). Participants could transition prematurely to the next phase while maintaining blinding, if requested by the participant or if deemed clinically necessary by the treating psychiatrist or research team, provided the HAM-D-17 score was at least 15. Medication and DBS settings remained unchanged throughout the crossover phase. Participants underwent MRI scans following both active and sham stimulation sessions.

The severity of depressive symptoms was evaluated at each assessment point (baseline, follow-up, active, sham) using the HAM-D-17, with higher scores indicating greater severity of depressive symptoms. Response to DBS was defined as a reduction of ≥ 50% in HAM-D-17 score at follow-up compared to baseline.

### Structural MRI data acquisition and analysis

#### Data acquisition

SMRI data were collected with a 1.5T Siemens Magnetom Avanto syngo MR scanner with a transmit/receive (Tx/Rx CP) Head Coil. A three dimensional single shot T1-weighted image with sagittal orientation was acquired (repetition time =1900 ms, echo-time =3.08 ms, flip angle =15^°^, matrix =512×512, number of slices =192, slice thickness =1.0mm, slice gap =50%, field of view =256×256, voxel-size =0.5×0.5×1.0 mm). For safety reasons, the DBS devices were switched off during scanning.

#### VBM Analysis

MRI data were preprocessed using the standardized CAT12 toolbox pipeline (r1450, http://www.neuro.uni-jena.de/cat) within SPM12 (v7487, https://www.fil.ion.ucl.ac.uk/spm/software/spm12) implemented in MATLAB (R2020a, The Mathworks, Natick, MA). This preprocessing involved correction for inhomogeneity, segmentation based on partial volume, and normalization to MNI space via Geodesic Shooting normalization (15), utilizing a template derived from 555 subjects of the IXI database (http://brain-development.org/) provided by the CAT12 toolbox. The resulting gray matter (GM) and white matter (WM) segmentations were modulated by the Jacobian determinant to account for volume changes during normalization. Quality control checks provided by the CAT12 toolbox, along with visual inspection, were employed to assess segmentation quality. Subsequently, the data underwent spatial smoothing using an 8 mm full-width-at-half-maximum kernel.

For VBM analysis, whole-brain GM and WM masks were generated by thresholding individual GM/WM images at 0.15 and retaining only voxels surviving thresholding across all patients. For the GM analysis, bilateral region-of-interest (ROI)-specific masks for the thalamus, amygdala, and hippocampus were created based on the Talaraich Daemon atlas using the WFU PickAtlas tool integrated into SPM12. Due to the large spatial extent of the prefrontal cortex and lack of a more regionally specific hypothesis, no ROI was constructed for the prefrontal cortex. For the WM analysis, bilateral ROI-specific masks for the anterior thalamic radiation (ATR) were extracted from the JHU white-matter tractography atlas (25% threshold of the maximum probability maps), which is included in the FSL library. This definition of the ATR also incorporates a significant portion of the superolateral medial forebrain bundle (slMFB) that runs in parallel to the ATR(16).

### Statistical analyses

#### Clinical and demographic data

We summarized clinical and demographic data of the entire sample. To investigate whether responders and non-responders differed on demographic variables at baseline and follow-up (symptom severity) we used t-tests and X^2^ -tests as appropriate. Tests were performed using SPSS (version 26).

#### Structural MRI analyses

All comparisons between groups, including TRD patients (n=12) versus healthy controls (n=16), responders (n=7) versus non-responders (n=5), were conducted at both ROI and whole-brain levels. Preprocessed and masked volume maps were utilized for these analyses. Baseline HAM-D-17 scores, age at baseline, sex, and total intracranial volume (TIV) were included as covariates in the analysis after demeaning. Voxel-wise statistical tests across the whole brain were family-wise error (FWE) rate corrected for multiple comparisons using threshold-free cluster enhancement (TFCE) with 10,000 permutations (17). Voxel-wise statistical tests within the combined ROI masks were FWE corrected at the peak-level using a small volume correction (SVC) (18).

In addition to group comparisons, regression analyses were performed between ROI/whole-brain segmentations and post-treatment HAM-D-17 scores, utilizing the same covariates and statistical procedures as outlined above.

## Results

In total, twenty-five patients and twenty-two healthy controls were included. Complete baseline and follow-up sMRI data were available for twelve patients and seventeen healthy controls (for the reasons for missing data, see Supplementary Table 2). Data of one healthy control was excluded due to excessive motion. Hence, the final sample for the baseline/follow-up analyses consisted of twelve patients (7 responders/5 non-responders) and sixteen healthy controls. Response status for one patient was manually adjusted from non-responder to responder as the patient had been in remission over the course of the optimization and the follow-up HAM-D-17 score was inflated by depression-unrelated physical complaints. Complete sMRI data during the cross-over phase were available for thirteen patients (7 responders/ 6 non-responders; for the reasons for missing data, see Supplementary Table 2).

### Clinical Results

Demographic and clinical characteristics are shown in Table 1. Patients and healthy controls did not differ significantly in sex, age and estimated IQ. As expected, patients scored significantly higher on baseline measures of depressive symptom severity scores (HAM-D-17, MADRS and IDS-SR). There was a significant reduction in HAM-D-17 score from baseline to follow-up (baseline: mean = 23.17, 95%CI = 19.39–26.94; follow-up: mean = 14.58, 95%CI = 9.03–20.13; t = 3.149, p =.009), and patients had a significantly lower HAM-D-score after active stimulation compared to sham stimulation (active: mean =14.92, 95%CI = 10.62–19.23; sham: mean = 22.54, 95%CI = 19.46–25.54; t = −3.200, p =.008).

**Table 1.** Descriptive characteristics of DBS patients and healthy controls. DBS Deep Brain Stimulation, HC healthy controls, SD standard deviation, IQ intelligence quotient, HAM-D Hamilton Rating Scale for Depression, MADRS Montgomery-Åsberg Depression Rating Scale, IDS-SR Inventory of Depressive Symptomatology–Self-report, MDD major depressive disorder, ECT electroconvulsive therapy. a χ^2^ test b Independent samples t-test c Paired t-test *Significant

| Characteristics | Baseline - follow-up |  | p-value |
| --- | --- | --- | --- |
|  | DBS | HC |  |
| Sample Size | 12 | 16 |  |
| Sex, No. (%) |  |  |  |
| Female | 9 (75.00) | 10 (62.50) | 0.687<sup>a</sup> |
| Male | 3 (25.00) | 6 (37.50) |  |
| Age at inclusion, mean (SD), y | 48.83 (2.59) | 54.00 (2.21) | 0.141<sup>b</sup> |
| Estimated IQ, mean (SD) | 93.08 (4.88) | 102.56 (3.48) | 0.116<sup>b</sup> |
| Baseline HAM-D-17 score, mean (SD) | 23.17 (1.71) | 1.13 (0.34) | < 0.001<sup>c, *</sup> |
| Baseline MADRS score, mean (SD) | 32.75 (3.13) | 1.31 (0.38) | < 0.001<sup>c, *</sup> |
| Baseline IDS-SR score, mean (SD) | 49.92 (4.37) | 3.25 (0.69) | < 0.001<sup>c, *</sup> |
| Age at MDD onset, mean (SD), y |  |  |  |
| Self-report | 25.00 (4.36) |  |  |
| Diagnosis | 34.00 (2.88) |  |  |
| No. of past medications, mean (SD) | 10.83 (1.05) |  |  |
| No. of past ECT series, mean (SD) | 2.00 (0.30) |  |  |
| No. of past ECT sessions, mean (SD) | 52.92 (15.12) |  |  |
| Years since MDD onset, diagnosis, mean (SD) | 14.83 (2.30) |  |  |
| MDD episodes, No. (%) |  |  |  |
| 1 | 6 (50.00) |  |  |
| 2 | 2 (16.67) |  |  |
| >2 | 4 (33.33) |  |  |

### Gray Matter Volume (GMV)

#### Baseline vs. Follow-up

Whole-brain comparisons between all patients and healthy controls showed the anticipated volume reduction at the electrode location, which is due to the implantation and signal distortion surrounding the electrodes (p_FWE_ < 0.05; peak voxel (x,y,z): [12,12,-8]) (See Supplementary Figure. 1). Analysis restricted to the ROI consisting of the thalamus, amygdala and hippocampus revealed a significant group x session interaction in the left anterior thalamus ([−2, −6, 10], p_SVC_ = 0.039), with a relative reduction over time in TRD patients compared to controls (Fig. 1A). This effect was primarily located in the left medial nucleus (Fig. 1A). Post-hoc comparisons however showed no significant difference in volume at baseline or follow-up between the two groups, and no significant change in thalamus, amygdala or hippocampus volume at follow-up compared to baseline within both groups (Fig. 1B-E).

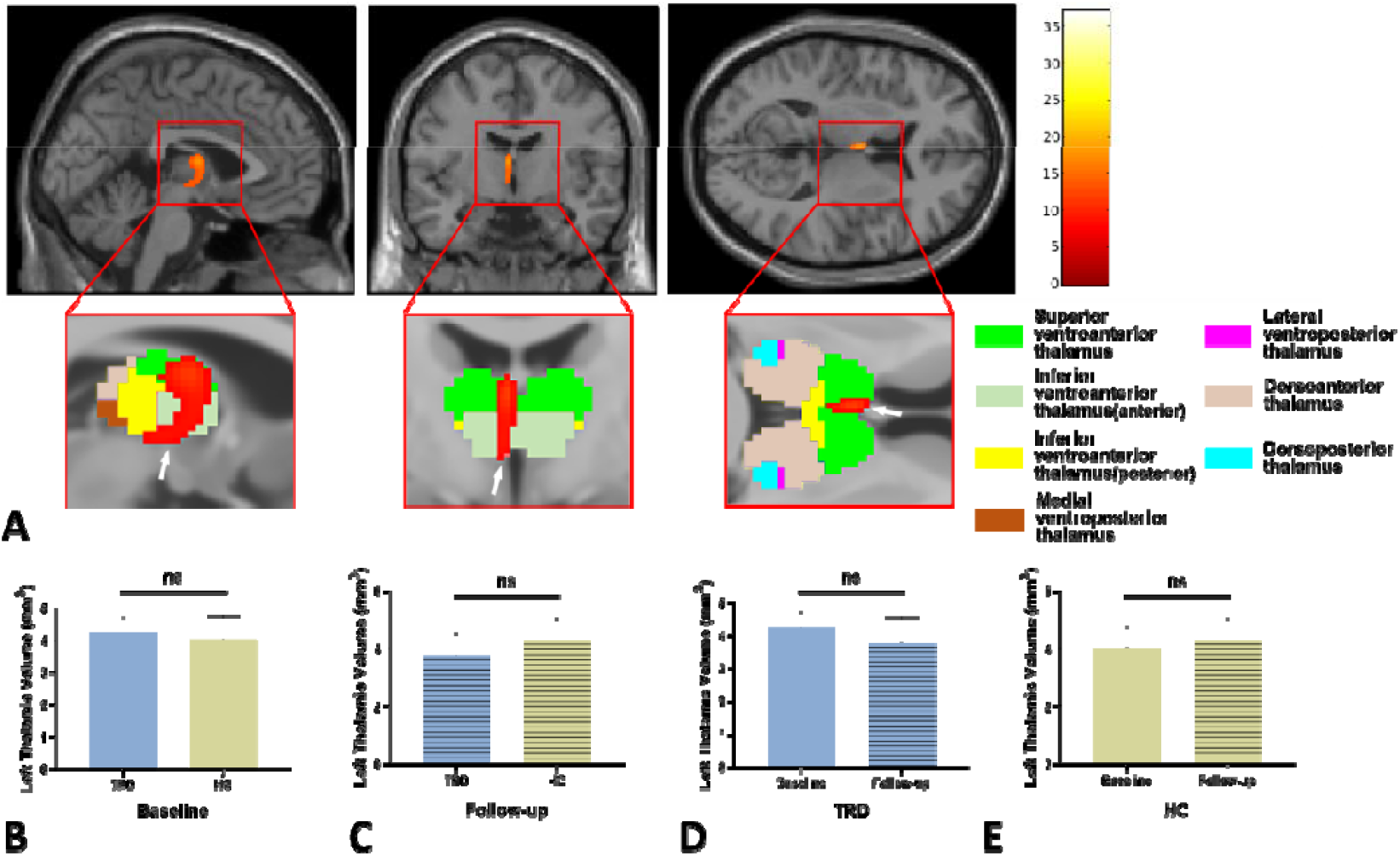

For exploratory purposes we divided patients into responder and non-responder groups and repeated the same analyses. Whole-brain comparison between these two groups did not yield any significant clusters (p_FWE_ > 0.05). The ROI-analysis showed a significant group x session interaction in the left posterior thalamus extending to the hippocampus ([−16, −34, 2], p_SVC_ = 0.014) (Fig. 2A). This effect was located in the dorsolateral nucleus of left thalamus (Fig. 2A). Post-hoc comparisons revealed a significant reduction in left thalamic volume at follow-up compared to baseline in non-responders (p <0.01) (Fig. 2E) but not in responders (Fig. 2D). Additionally, no significant difference in volume was found at baseline or follow-up between the two groups (Fig. 2B-C).

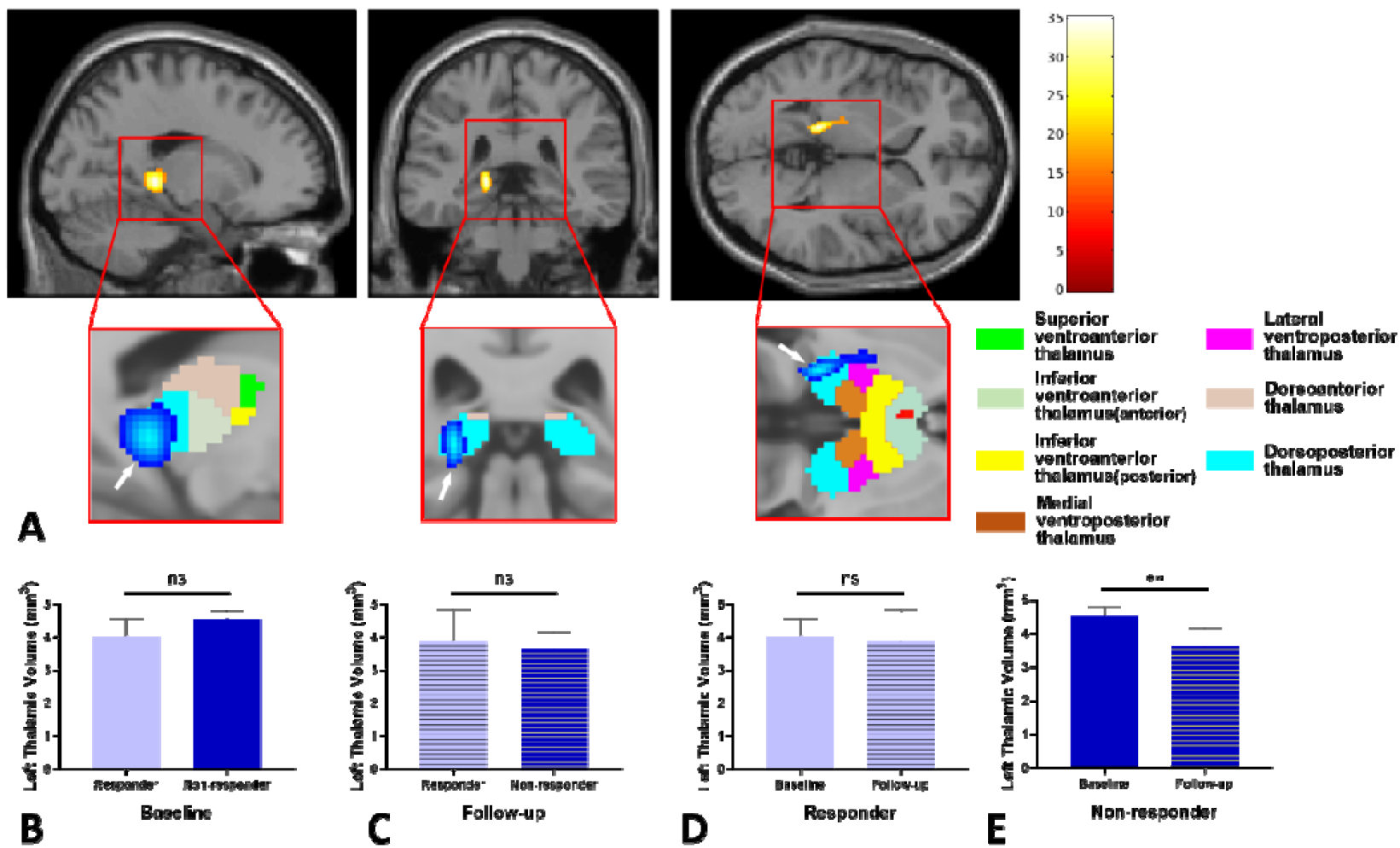

We then compared responders or non-responders to healthy controls separately. Both whole-brain analysis revealed the significant volume change at the electrode location [12,12, 8]. The ROI analysis did not show a significant difference in GMV change in responders compared to healthy controls. But for non-responders compared to healthy controls, the ROI analysis showed a significant group x session interaction in the left thalamus ([−2, −6 10], p_SVC_ = 0.001) (Fig. 3A). Post-hoc comparison revealed larger left thalamic volume in non-responders than healthy controls at baseline (p<0.01) (Fig. 3B), but no significant differences at follow-up (Fig. 4C). In addition, left thalamic volume changed at follow-up compared to baseline in non-responders (p< 0.01) (Fig. 3D) but not in healthy controls (Fig. 3E).

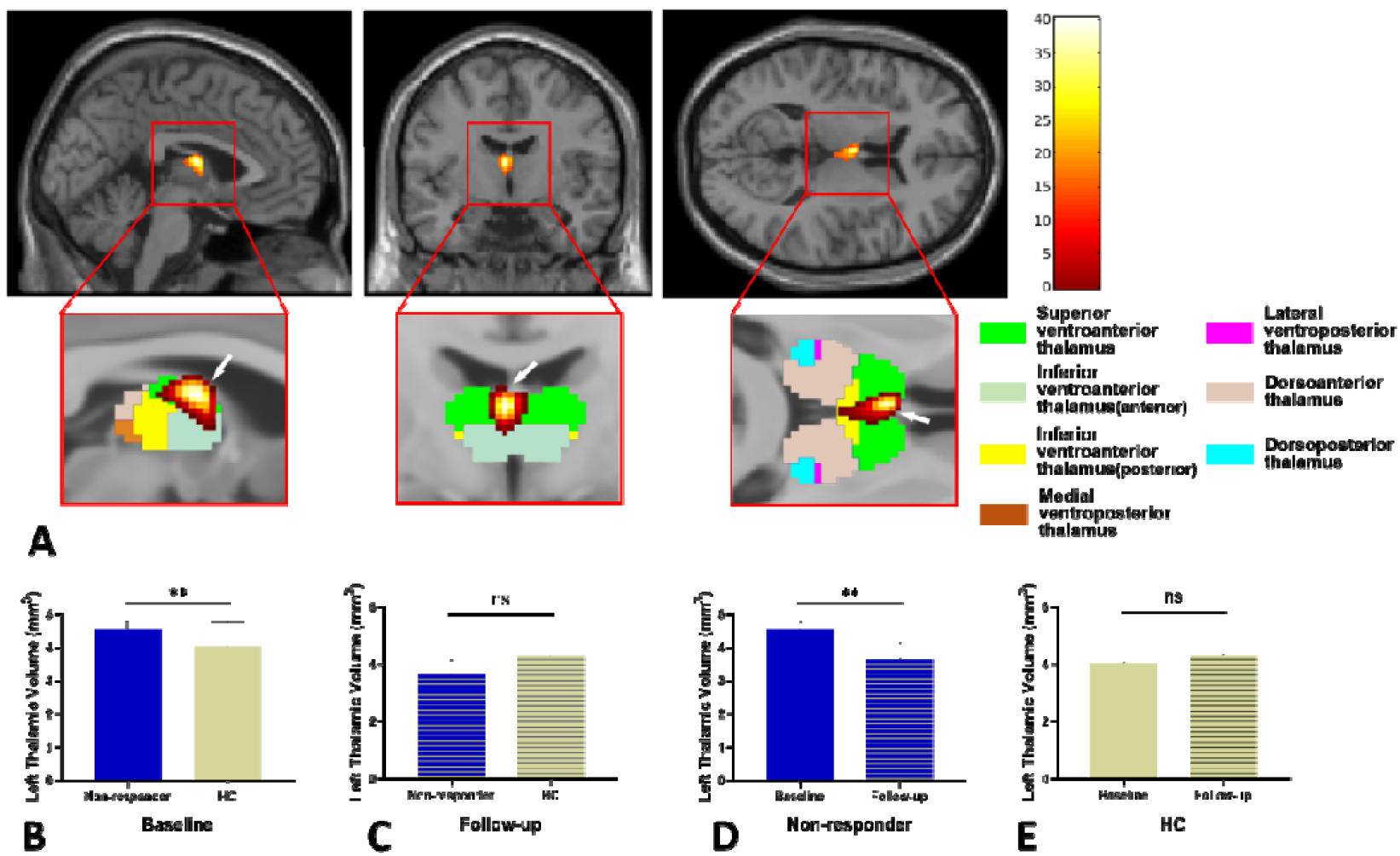

#### Active vs. sham DBS

Whole-brain analysis and ROI analysis in the cross-over phase (active vs. sham) showed no significant differences in GMV change.

### White Matter Volume (WMV)

#### Baseline vs. Follow-up

Whole-brain comparisons between all patients and healthy controls revealed significant WMV changes along the surgical pathway (p_FWE_ < 0.05; See Supplementary Figure. 2), but ROI analysis within the ATR ROI did not yield any significant clusters (p_FWE-corrected_ > 0.05).

#### Active vs. sham DBS

Whole-brain analysis and ATR-ROI analysis in the cross-over phase (active vs. sham) showed no significant differences in any specific brain region.

## Discussion

The aim of present study was to elucidate volumetric brain alterations in TRD patients following vALIC DBS treatment. We found that long-term vALIC DBS reduced left anterior thalamic gray matter volume in TRD patients in comparison to healthy controls. Exploratory follow-up analyses suggested that this might be explained by larger thalamus volumes at baseline in patients who did not respond to DBS compared to HC, which had decreased more at follow-up than in responders as well as HC. No significant changes in WMV were observed. Short-term DBS cessation during the sham-controlled crossover phase did not show significant effects. To our knowledge, this is the first study demonstrating structural plasticity after long-term DBS in TRD.

A limited number of studies have investigated DBS-related neuroanatomical changes in other disorders, and have also noted thalamic volume alterations. Notably, DBS induces comparable changes in neurological disorders, including a reduction in thalamic volume in patients with Parkinson’s disease (19-21). These studies did not only observe changes in the ipsilateral thalamus, but also in other brain regions outside the surgical pathway, including the putamen, hippocampus, and even alterations in other cortical regions and entire hemispheres, indicating that DBS treatment induces changes in brain volume beyond the target area. The volume reduction after vALIC DBS in our study thus appears rather specific, though this may turn out to be more extensive in larger studies with more statistical power. And whereas thalamic volume decreases after DBS, it increases after electroconvulsive therapy (ECT) (22, 23). Interestingly, the increase in thalamic volume seems temporary, as it decreases again after the cessation of ECT in the 10-36 months thereafter, especially in ECT responders (24). This suggests that the brain of depressed patients exhibits plasticity, and that it requires longer and persistent modulation to manifest. We did not observe changes in thalamic volume during the cross-over phase, despite the short-term symptomatic improvements observed with active compared to sham stimulation.

The observed volumetric alteration across all TRD patients occurred in the anterior medial nuclear clusters of the left thalamus. The thalamus is a complex structure composed of various subunits, including the anterior, lateral, ventral, intralaminar, medial, and posterior parts (25). Each subunit forms connections with various brain structures, including extensive connections with the hippocampus and limbic cortical regions (26, 27). Recent research has revealed diverse structural differences within specific thalamic subregions across various mental diseases (28, 29). Since the target of this study was the ALIC, we focused more on the connections between different thalamic subnuclei and the ALIC. The ALIC contains projections from the mediodorsal thalamus pars magnocellularis, which then connects to the orbitofrontal cortex (30). The mediodorsal thalamus, along with several other brain regions, comprises several large-scale neuronal networks implicated in mediating the emotional and cognitive symptoms of depression (31, 32), which may thus also be affected by vALIC DBS. The comparison between responders and non-responders revealed group differences in the posterior dorsolateral subregion. We speculate that the volume alteration in the dorsolateral nuclei that was related to DBS response may be due to its connection with the limbic system (33). This may in part be mediated by changes in the CSTC circuit, as there currently is no evidence to suggest a direct connection between the ALIC and the dorsolateral thalamus.

Exploratory analysis suggested pretreatment thalamic volume differences between TRD patients and healthy controls. TRD patients who did not respond to vALIC DBS had a larger thalamic volume while responders exhibited no significant differences in thalamic volume compared to healthy controls. Several previous studies have indicated a relationship between thalamic volume and the severity of depressive symptoms (34, 35), as well as its association with treatment response. Larger pre-treatment thalamic volume is predictive of poor response to ECT(36), but it has also been associated with good response after naturalistic follow-up(37). And one DBS study reported that preoperative thalamic volumes were significantly larger in patients who responded to SCC-DBS compared to non-responders, but this effect was not significant when accounting for differences in brain size (38). In contrast, our study showed larger left thalamic volumes in patients with a poor DBS response. This suggests that thalamic abnormalities impede the response to DBS, which needs to be replicated in future studies.

We also investigated whether vALIC DBS altered WMV in the present study. A series of previous studies had confirmed white matter volumetric differences in MDD patients. (39), (40). (38). But aside from changes in WMV along the surgical pathway, we did not find any other significant alterations, whether in comparisons between TRD patients and healthy controls at baseline or between preoperative and postoperative patient groups. We therefore speculate that vALIC DBS may primarily induce gray matter rather than white matter plasticity, even though the electrodes are implanted in white matter. The ALIC does play an important role in the pathophysiology of depression, as both the ATR and slMFB have been implicated in depression (6, 8, 41, 42). Compared to healthy controls, adults with severe MDD exhibit reduced white matter fractional anisotropy (FA) in the ALIC, with FA showing a negative correlation with symptom severity (43, 44). Surgical interventions for MDD further confirm the association between ALIC and depression, as bilateral anterior capsulotomy has been employed for TRD (45, 46). But as we could not investigate white matter changes near the electrodes due to signal dropout, we cannot exclude the possibility that DBS affects the white matter near the DBS contact points.

The main limitation of this study is the small sample size. This is inherent to studies of DBS in TRD, as DBS is not yet a standard treatment for depression and clinical trials are also restricted to small samples. Additionally, the patients in this study were still receiving antidepressant treatment besides DBS, which might have influenced brain structure. But as patients were already taking medication before the start of DBS, and since there was no significant volumetric difference between all patients and healthy individuals at baseline, we presume that the effects of antidepressant medication were limited.

In conclusion, these results show that long-term vALIC DBS is associated with a reduction in anterior thalamus GMV in patients with TRD. No changes in WMV or after short-term stimulation cessation were observed. Together, these results suggest that long-term DBS might result in structural plasticity of the thalamus.

## Data Availability

The code used in the current study is available upon request. Raw data cannot be shared due to IRB and informed consent restrictions.

