## Supplemental Data for "Structural plasticity following deep brain stimulation of the internal capsule in treatment resistant depression"

Abstract

Objective

Deep brain stimulation (DBS) targeting the ventral anterior limb of the internal capsule (vALIC) shows potential as treatment for treatment resistant depression (TRD). While DBS alters brain function, it is not yet known whether it also induces anatomical alterations. Here we investigated the long-term effects of vALIC DBS on brain structure using structural magnetic resonance imaging.

Methods

We included data from twelve patients with TRD before DBS surgery and after DBS parameter optimization, and from sixteen matched healthy controls at a comparable time interval to control for test-retest effects. To investigate the short-term effects of DBS deactivation after parameter optimization, thirteen patients were additionally scanned after double-blind periods of active and sham stimulation. Voxel-based morphometry analysis was used to identify volumetric differences.

Result

The group x time interaction showed significant changes in anterior thalamic gray matter volume, with a relative reduction in TRD patients compared to controls. Follow-up analysis suggested that this was related to larger thalamic volume at baseline in DBS non-responders. A direct comparison between responders and non-responders showed an additional interaction in the posterior thalamus and hippocampus, which was also related to larger volumes at baseline in non-responders. The comparison between active and sham stimulation during the crossover phase did not show significant differences.

Conclusion

These results show that long-term vALIC DBS is associated with a reduction in thalamic volume compared to healthy controls, suggesting that long-term DBS induces focal structural plasticity.

Key words

Treatment Resistant Depression, Internal Capsule, Deep Brain Stimulation, Voxel-based morphometry

**Supplementary Data**

1.Supplementary Methods

1.1 Clinical and behavioral statistical analyses

Clinical data were analyzed with R version 4.0.3 (R Core Team,2020).HAM-D-17 scores at baseline and follow-up were compared with a linear mixed-effects model analysis with HAM-D-17 score as the dependent variable and session (baseline vs.follow-up) and time between sessions as independent variables.In addition,for the active/sham stimulation comparison we performed a linear mixed-effects model analysis with HAM-D-17 as the dependent variable and session (active vs.sham),time between sessions,and randomization order as independent variables.

1.2 MRI Acquisition Parameters

A three dimensional single shot T1-weighted image with sagittal orientation was acquired (repetition time=1900 ms,echo-time=3.08 ms,flip angle=15°, matrix=512x512,number of slices=192,slice thickness=1.0 mm,slice gap=50%,field of view=256x256,voxel-size=0.5x0.5x1.0 mm).

2. Supplementary Table

Supplementary Table 1. Number of patients using psychotropic medication overtime

|  |  | **Baseline - follow-up (n=12)** | | **Active-sham (n=13)** | |
| --- | --- | --- | --- | --- | --- |
|  |  | **Baseline** | **Follow-up** | **Baseline** | **Follow-up** |
| **Antidepressant** | Combination | 1 | 1 | 1 | 3 |
|  | Single | 6 | 4 | 7 | 4 |
|  | None | 5 | 7 | 5 | 6 |
| **Benzodiazepine** | Combination | 1 | 0 | 0 | 0 |
|  | Single | 4 | 5 | 5 | 5 |
|  | None | 7 | 7 | 8 | 8 |
| **Antipsychotic** | Single | 5 | 5 | 6 | 5 |
|  | None | 7 | 7 | 7 | 8 |
| **Lithium** | Single | 1 | 0 | 1 | 1 |
|  | None | 11 | 12 | 12 | 12 |
| **Anxiolytic** | Single | 0 | 1 | 0 | 1 |
|  | None | 12 | 11 | 13 | 12 |
| **Anti-epileptic** | Single | 1 | 0 | 1 | 1 |
|  | None | 11 | 12 | 12 | 12 |
| **Antihistaminic** | Single | 1 | 1 | 1 | 1 |
|  | None | 11 | 11 | 12 | 12 |
| **Opioid** | Single | 0 | 0 | 1 | 1 |
|  | None | 12 | 12 | 12 | 12 |
| **Sympathomimetic** | Single | 1 | 1 | 1 | 1 |
|  | None | 11 | 11 | 12 | 12 |

Supplementary Table 2.Reasons for missing MRI data.

| **Patient** | **Complete** **baseline/** **follow-up** **MRI** **data** | **Complete** **cross-over** **phase** **MRI** **data** | **Reason** **for** **missing** **data** |
| --- | --- | --- | --- |
| 1 | **No** | **No** | MRI coil was unavailable at baseline.Because there was no baseline data it was decided to not collect data at follow-up and during the cross-over phase. However,from this patient onwards MRI data was collected at the following  assessments despite missing baseline data |
| 2 | No | Yes | MRI coil was unavailable at baseline. |
| 3 | No | Yes | MRI coil was unavailable at baseline. |
| 4 | Yes | Yes |  |
| 5 | Yes | No | Patient was deemed unfit to participate in the cross-over phase due to unstable clinical status |
| 6 | No | No | Drop-out due to non-response. |
| 7 | No | No | Patient was treated with MRI-incompatible vagus nerve stimulation. |
| 8 | No | No | Drop-out due to non-response. |
| 9 | Yes | Yes |  |
| 10 | No | No | Drop-out due to non-response |
| 11 | Yes | Yes |  |
| 12 | No | No | Follow-up time deviated too much from protocol (2.5 years).Patient was deemed unfit to participate in the cross-over phase due to unstable clinical status. |
| 13 | Yes | Yes |  |
| 14 | No | No | Patient withdrew from participation after the baseline assessment due to somatic complaints |
| 15 | Yes | Yes |  |
| 16 | No | No | Drop-out due to non-response |
| 17 | Yes | Yes |  |
| 18 | Yes | No | Unknown |
| 19 | No | Yes | Baseline data was not collected due to back complaints at the time. |
| 20 | Yes | No | MRI data collection was terminated due to an anxiety attack during one of the cross-over assessments |
| 21 | Yes | Yes |  |
| 22 | No | Yes | Unknown |
| 23 | No | Yes | MRI data collection was terminated due to an anxiety attack at the follow-up assessment |
| 24 | Yes | Yes |  |
| 25 | Yes | No | Patient was deemed unfit to participate in the cross-over phase due to unstable clinical status |
| **Healthy** **control** | **Complete** **baseline/** **follow-up** **MRI** **data** |  | **Reason** **for** **missing** **data** |
| 1 | Yes |  |  |
| 2 | Yes |  |  |
| 3 | Yes |  |  |
| 4 | Yes |  |  |
| 5 | Yes |  |  |
| 6 | Yes |  |  |
| 7 | Yes |  |  |
| 8 | No |  | Participant withdrew from participation after the baseline assessment |
| 9 | Yes |  |  |
| 10 | Yes |  |  |
| 11 | Yes |  |  |
| 12 | Yes |  |  |
| 13 | No |  | Unknown |
| 14 | Yes |  |  |
| 15 | No |  | Unknown |
| 16 | No |  | MRI-incompatible elbow pin |
| 17 | No |  | MRI data collection was terminated due to an anxiety attack |
| 18 | Yes |  |  |
| 19 | Yes |  |  |
| 20 | Yes |  |  |
| 21 | Yes |  |  |
| 22 | Yes |  |  |

Abbreviations:MRI,magnetic resonance imaging

3. Supplementary Figure

Supplementary Figure 1. Whole-brain comparisons between all patients and healthy controls showed the significant gray matter volume change in the electrode location[12,12,-8].

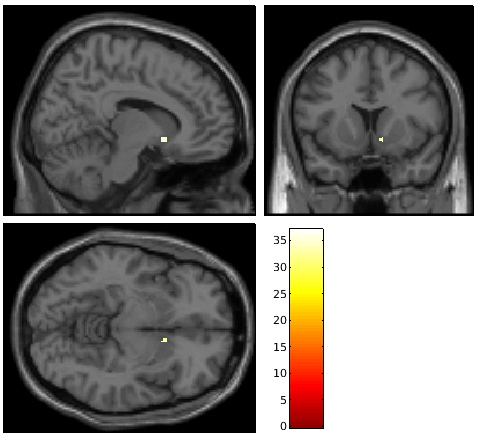

Supplementary Figure 2. Whole-brain comparisons between all patients and healthy controls showed the significant white matter volume change along the surgical pathway.

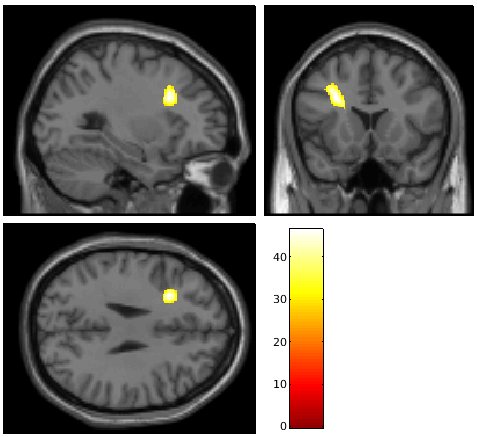
